# Behavioural change towards reduced intensity physical activity is disproportionately prevalent among adults with serious health issues or self-perception of high risk during the UK COVID-19 lockdown

**DOI:** 10.1101/2020.05.12.20098921

**Authors:** Nina Trivedy Rogers, Naomi Waterlow, Hannah Brindle, Luisa Enria, Rosalind M Eggo, Shelley Lees, Chrissy h Roberts

**Affiliations:** London School of Hygiene and Tropical Medicine, London WC1E 7HT, UK; UCL Research Department of Epidemiology & Public Health, London WC1E 7HB, UK; University of Bath, Department of Social & Policy Sciences, Bath BA2 7AY

## Abstract

**Importance:** There are growing concerns that the UK COVID-19 lockdown has reduced opportunities to maintain health through physical activity, placing individuals at higher risk of chronic disease and leaving them more vulnerable to severe sequelae of COVID-19.

**Objective:** To examine whether the UK’s lockdown measures have had disproportionate impacts on intensity of physical activity in groups who are, or who perceive themselves to be, at heightened risk from COVID-19.

**Designs, Setting, Participants:** UK-wide survey of adults aged over 20, data collected between 2020-04-06 and 2020-04-22.

**Exposures:** Self-reported doctor-diagnosed obesity, hypertension, type I/II diabetes, lung disease, cancer, stroke, heart disease. Self-reported disabilities and depression. Sex, gender, educational qualifications, household income, caring for school-age children. Narrative data on coping strategies.

**Main Outcomes and Measures:** Change in physical activity intensity after implementation of UK COVID-19 lockdown (self-reported).

**Results:** Most (60%) participants achieved the same level of intensity of physical activity during the lockdown as before the epidemic. Doing less intensive physical activity during the lockdown was associated with obesity (OR 1.21, 95% CI 1.02-1.41), hypertension (OR 1.52, 1.33-1.71), lung disease (OR 1.31,1.13-1.49), depression (OR 2.02, 1.82-2.22) and disability (OR 2.34, 1.99-2.69). Participants who reduced their physical activity intensity also had higher odds of being female, living alone or having no garden, and more commonly expressed sentiments about personal or household risks in narratives on coping.

**Conclusions and relevance:** Groups who reduced physical activity intensity included disproportionate numbers of people with either heightened objective clinical risks or greater tendency to express subjective perceptions of risk. Policy on exercise for health during lockdowns should include strategies to facilitate health promoting levels of physical activity in vulnerable groups, including those with both objective and subjective risks.

## Introduction

The pandemic spread of Severe Acute Respiratory Syndrome Coronavirus 2 (SARS-CoV-2) [1] was declared a Public Health Emergency of International Concern by the World Health Organisation on 30 January 2020 [2] and by the end of April 2020 the virus had infected more than 3 million people worldwide, causing more than 200,000 deaths [3]. In order to limit the spread of COVID-19, governments across the globe imposed varying degrees of social distancing advice and nationwide lockdowns. On 23 March 2020 the UK government enacted measures that were included in the Coronavirus Act 2020 and recommended that everyone must stay in their homes unless (i) shopping for essentials such as food and medicine, (ii) requiring medical assistance, (iii) caring for vulnerable people, (iv) travelling to and from work if absolutely necessary and (v) to carry out one form of exercise (e.g: walking, running, cycling) each day, either alone or with people who live together. Some adults aged 70 and over and those with specific underlying health conditions including asthma, heart disease, diabetes and being seriously overweight were also advised to follow much stricter social isolation recommendations. In this paper we refer to the combined package of measures as ‘lockdown’.

There have been growing concerns that the limitations lockdown has placed on opportunities for individuals to be physically active could have public health implications [4,5], The tradeoff between protection from COVID-19 and increased risk of inactivity presents already vulnerable populations with a potential “no-win” situation; for instance where the consequence of protection from acquiring SARS-CoV-2 infection is increased inactivity, which could put these same individuals at heightened risk of mental health problems [6], chronic diseases, such as cardiovascular disease, stroke [7,8] and premature mortality [9], Longer term, it is therefore also possible that because of lockdown-associated increases in underlying health conditions [10], the effects of changes in physical activity (PA) during lockdown could actually serve to increase the size of the population that is vulnerable to severe complications from COVID-19 in subsequent epidemic waves. Reduced PA may also have a negative impact on the control of chronic health problems including metabolic, cardiovascular, musculoskeletal, pulmonary and psychiatric conditions; all of which are often better controlled when PA is included as part of the management plan [11], These effects could potentially add additional pressure to the health system during the current or later epidemic waves.

The WHO and UK guidelines on PA for adults recommend at least 150 minutes of moderate PA, 75 minutes of vigorous PA, or some equivalent combination of the two per week [12], Newly revised guidelines by the UK Chief Medical Officers also emphasised that some PA is better than none and that even light activity brings some health benefits compared to being inactive, especially in the case of older adults who are more likely to live with chronic health conditions [13],

In this study we identify whether the UK’s lockdown measures have had disproportionate impacts on PA intensity in groups who are, or who perceive themselves to be at risk of worse outcomes of COVID-19 disease. This study takes the form of a UK-wide survey of adults aged over 20.

## Methods

### Online Survey

Anonymous survey data were collected online between 2020-04-06 and 2020-04-22, roughly mapping to weeks 3-5 of the lockdown in the UK. The survey included 49 questions which covered a broad range of topics including (1) Demographics, (2) Health and Health Behaviours, (3) Adherence to COVID-19 Control measures, (4) Information sources used to learn about COVID-19, (5) Trust in various information sources, government and government decision-making, (6) Rumours and misinformation, (7) Contact & Communication during COVID-19 and (8) Fear and Isolation.

The survey was publicised using a ‘daisy-chaining’ approach in which respondents were asked to share and to encourage onward sharing of the survey’s Uniform Resource Locator (URL) among friends & colleagues. The study team directly targeted a number of faith institutions, schools and special interest groups and also used Facebook’s premium “Boost Post” feature. A “boosted” post functions as an advert which can be targeted at specific demographics. We boosted details of the survey and it’s URL to a target audience of 113,280 Facebook users aged 13-65+ years and living in England, Wales, Scotland and Northern Ireland. Participants were also provided with URL links to a set of freely available summary reports and analyses which were periodically updated in near-real time.

We used an ODK XForm (https://eetodk.github.io/xforms-spec/) deployed on Enketo smart paper (https://enketo.org/) via ODK Aggregate v.2.0.3 (https://github.com/getodk/aggregate). Form level encryption and end-to-end encryption of data transfer were implemented on all submissions.

### Disability and classification of health conditions

Participants were assessed for disability by asking about difficulties in six activities of daily living (ADLs) [14] including bathing, dressing, walking across a room, eating (such as cutting up food), getting in and out of bed, and using the toilet (including getting up and down). Disability was defined by the presence of at least one ADL. We also explored depressive symptomatology with the question *“In the past two weeks, how often have you felt down, depressed, or hopeless?”*. Options were *“not at all”, “several days”, “more than half the days”* and *“every day”*. Participants were classified as currently depressed if they reported feeling this way either *“more than half the days”* or *“every day”*. To determine whether patients had any previous or current chronic disease(s) diagnosis (CDD), participants were asked *“Has a doctor ever diagnosed you with any of the following?”*. The question allowed for multiple chronic diseases to be selected from a list that included diabetes type 1, diabetes type 2, lung disease, cancer, stroke, heart disease, high blood pressure (hypertension) and obesity.

We additionally asked participants to provide (in narrative text form) details about any other medical conditions that they felt would increase their risk of getting seriously ill if they were to catch coronavirus. We chose to recode any participant who mentioned asthma as having a lung disease because the topic of *“Asthma”* accounted for around 25% of the open text responses to this question (Determined by structural text modelling, see below) and because asthma was mentioned directly by 678 participants (Supplementary Figure S1). The majority of people who reported having a doctor’s diagnosis of lung disease also mentioned asthma (63.4%, n = 225, Supplementary Table SI) suggesting that they operationalised asthma as a lung disease and may have been referring to asthma when they reported their prior diagnosis of lung disease. 8.3% (n = 453) of people who did not report having ‘lung disease’ did however mention asthma in the free text.

### Analysis

We performed a complete case analysis of male and female gendered participants aged 20 years and over, opting to include only participants who had provided responses to all the relevant fields including baseline PA, PA during lockdown, highest educational qualifications, age (20-34; 35-54; 55-69; 70+), gender, whether living alone, household income, presence of ADLs, self-rated depression and pre-existing chronic diseases. Pearson’s χ^2^ test was used to detect factor variables with statistically significant differences between the groups when the data were grouped according to baseline (pre-COVID-19) PA levels. Due to significant differences according to baseline PA, all. further analyses were corrected for baseline PA. The main response variable for statistical association tests was any change in PA intensity from pre-COVID-19 lockdown to the time of survey participation. This value was calculated by comparing baseline PA *(“Before the outbreak began, what type of exercise did you regularly do?”*, options *“None”, “Mild [e.g. walking short distances, doing DIY etc.]”, “Moderate [e.g. A gentle workout, Digging the garden, Dancing]”* & *“Vigorous [e.g. Running/Jogging/Hiking, Cycling, Weightlifting]”)* to PA during COVID-19 lockdown (*“What type of exercise are you doing now?”*, options as for baseline). Participants were classified as doing the “Same”, “Less” or “More” than their usual PA intensity. Using the ‘nnet’ R package, we applied a multinomial log-linear model via neural networks [15] to the detection of factors which were associated with change in PA intensity during lockdown.

### Topic Modelling

We used Structural Text Modelling (STM) [16] to identify key topics in the data on self-perceived medical risk factors (see above), and also to determine whether changes in PA intensity were associated with participants’ other perceptions of risk from COVID-19. STM employs machine learning (ML) approaches to explore open ended survey questions in a highly structured and reproducible way [16]. The goal of STM is to identify topics and perspectives in free-text data, for instance by highlighting specific diseases, themes or perspectives being reported in the survey. This is functionally analogous and equivalent in results to the type of human coding of text data performed by anthropologists and ethnographers; but unlike more conventional topic modelling, STM makes it possible to link topic models to metadata and quantitative data in a way that is directly amenable to statistical modelling [16,17], All STM was performed using the ‘stm’ package [17] for R. STM was applied to data from the open ended survey question “*On 23rd March 2020, the Prime Minister Boris Johnson announced a complete lockdown in the UK. Tell us what you have been doing to help you cope during this difficult time?’’*. The text data were processed into a corpus and transliterated to lower-case. Numbers, common punctuation and stop-words (such as *“I”, “me”, “that’s”* and *“because”*) were stripped and data were trimmed to include only words which appeared in 20 or more responses to the survey The corpus was then bound to the quantitative data from the survey and the STM was optimised to determine the number of topics which maintained the balance between high semantic coherence (i.e. the topics were clear and understandable) and exclusivity (vocabulary and themes had little cross-over between topics). The topics were then labelled manually (this and defining the number of topics of interest were the only subjective components of the process) by first examining the word usage within topics (weighted by exclusivity) and then assessing a number of representative perspectives (quotes) from each of the topics. Expected text proportions (ETP) were defined as the proportion of the total corpus which related to each topic. Between-topic correlations were measured using the semiparametric procedure described in the R package “huge” (High-Dimensional Undirected Graph Estimation). Tests for statistical associations between the PA data set and the STM topics used regressions of the STM, where the between group ETPs were the outcome variables and the survey PA question data, including the change in PA intensity were the explanatory variables.

### Patient and public involvement

This project uses tools and methods that have been developed as part of projects that were guided from the earliest stages by patient and public involvement and stakeholders have been included in all stages of the research. The open source survey software used in this study was developed in collaboration with a global community of researchers, data scientists and field epidemiologists, including members of the public, not-for-profit organisations and partners from low and middle income countries. A group of around 15 lay members of the UK public, including both younger and older people, were asked to test, review and recommend changes to the content of the survey before it was fully deployed.

### Ethics, Confidentiality & Participant wellbeing

The study was approved by the London School of Hygiene and Tropical Medicine Observational research ethics committee (Ref: 21846). All data were fully anonymous and the study team had no means by which they could identify individual respondents. All participants provided consent to participate in the study by ticking a box on the survey web-form. All questions in the survey were optional (excepting age and number of people in the household), meaning that participants could skip questions if they did not want to divulge specific data.

## Results

The survey consisted of 9,456 participants. After filtering the data (Supplementary Table S2) to a complete case analysis we retained 5,820 participants for analysis and demographic characteristics of the sample are given in Table 1.

**Table 1:** Demographic characteristics of the complete case sample, by baseline PA intensity

|  |  | PA Intensity before lockdown |  |  |  | Total (N=5820) | p value |
| --- | --- | --- | --- | --- | --- | --- | --- |
|  |  | none (N=168) | mild (N=1837) | moderate (N=2427) | vigorous (N=1388) |  |  |
| PA intensity during lockdown |  |  |  |  |  |  | < 0.001 |
|  | none | 89 (20.2%) | 234 (53.1%) | 82 (18.6%) | 36 (8.2%) | 441 (100.0%) |  |
|  | mild | 49 (2.5%) | 1126 (57.9%) | 603 (31.0%) | 168 (8.6%) | 1946 (100.0%) |  |
|  | moderate | 25 (1.0%) | 424 (17.5%) | 1616 (66.7%) | 356 (14.7%) | 2421 (100.0%) |  |
|  | vigorous | 5 (0.5%) | 53 (5.2%) | 126 (12.5%) | 828 (81.8%) | 1012 (100.0%) |  |
| PA Change during lockdown |  |  |  |  |  |  | < 0.001 |
|  | Same | 89 (2.4%) | 1126 (30.8%) | 1616 (44.2%) | 828 (22.6%) | 3659 (100.0%) |  |
|  | less | 0 (0.0%) | 234 (15.8%) | 685 (46.3%) | 560 (37.9%) | 1479 (100.0%) |  |
|  | more | 79 (11.6%) | 477 (69.9%) | 126 (18.5%) | 0 (0.0%) | 682 (100.0%) |  |
| Age |  |  |  |  |  |  | < 0.001 |
|  | 20-34 | 17 (3.4%) | 123 (24.6%) | 158 (31.6%) | 202 (40.4%) | 500 (100.0%) |  |
|  | 35-54 | 72 (3.0%) | 740 (31.3%) | 878 (37.2%) | 673 (28.5%) | 2363 (100.0%) |  |
|  | 55-69 | 69 (2.8%) | 784 (31.8%) | 1153 (46.7%) | 461 (18.7%) | 2467 (100.0%) |  |
|  | 70+ | 10 (2.0%) | 190 (38.8%) | 238 (48.6%) | 52 (10.6%) | 490 (100.0%) |  |
| Gender |  |  |  |  |  |  | < 0.001 |
|  | female | 133 (2.9%) | 1472 (32.4%) | 1973 (43.4%) | 965 (21.2%) | 4543 (100.0%) |  |
|  | male | 35 (2.7%) | 365 (28.6%) | 454 (35.6%) | 423 (33.1%) | 1277 (100.0%) |  |
| Living alone |  |  |  |  |  |  | 0.031 |
|  | no | 134 (2.7%) | 1513 (31.0%) | 2042 (41.9%) | 1190 (24.4%) | 4879 (100.0%) |  |
|  | yes | 34 (3.6%) | 324 (34.4%) | 385 (40.9%) | 198 (21.0%) | 941 (100.0%) |  |
| Education |  |  |  |  |  |  | < 0.001 |
|  | primary | 23 (4.6%) | 209 (41.9%) | 200 (40.1%) | 67 (13.4%) | 499 (100.0%) |  |
|  | alevel | 57 (3.7%) | 551 (35.6%) | 663 (42.8%) | 278 (17.9%) | 1549 (100.0%) |  |
|  | higher | 88 (2.3%) | 1077 (28.6%) | 1564 (41.5%) | 1043 (27.7%) | 3772 (100.0%) |  |
| Access to a garden |  |  |  |  |  |  | < 0.001 |
|  | yes | 145 (2.8%) | 1644 (31.3%) | 2252 (42.9%) | 1207 (23.0%) | 5248 (100.0%) |  |
|  | no | 23 (4.0%) | 193 (33.7%) | 175 (30.6%) | 181 (31.6%) | 572 (100.0%) |  |
| School aged children |  |  |  |  |  |  | < 0.001 |
|  | no | 122 (2.8%) | 1433 (32.3%) | 1897 (42.8%) | 983 (22.2%) | 4435 (100.0%) |  |
|  | yes | 46 (3.3%) | 404 (29.2%) | 530 (38.3%) | 405 (29.2%) | 1385 (100.0%) |  |
| Income |  |  |  |  |  |  | < 0.001 |
|  | Less than £15,000 | 38 (5.7%) | 262 (39.6%) | 287 (43.4%) | 75 (11.3%) | 662 (100.0%) |  |
|  | £15,000 - £24,999 | 21 (2.1%) | 374 (37.4%) | 437 (43.7%) | 169 (16.9%) | 1001 (100.0%) |  |
|  | £25,000 - £39,999 | 38 (2.9%) | 418 (32.4%) | 562 (43.5%) | 274 (21.2%) | 1292 (100.0%) |  |
|  | £40,000 - £59,999 | 41 (3.2%) | 362 (28.6%) | 523 (41.4%) | 338 (26.7%) | 1264 (100.0%) |  |
|  | £60,000 - £99,999 | 21 (1.9%) | 298 (27.3%) | 438 (40.1%) | 336 (30.7%) | 1093 (100.0%) |  |
|  | More than £100,000 | 9 (1.8%) | 123 (24.2%) | 180 (35.4%) | 196 (38.6%) | 508 (100.0%) |  |
| Disability (ADL) |  |  |  |  |  |  | < 0.001 |
|  | no | 134 (2.4%) | 1715 (30.6%) | 2387 (42.5%) | 1377 (24.5%) | 5613 (100.0%) |  |
|  | yes | 34 (16.4%) | 122 (58.9%) | 40 (19.3%) | 11 (5.3%) | 207 (100.0%) |  |
| Depression |  |  |  |  |  |  | 0.039 |
|  | no | 147 (2.8%) | 1631 (31.1%) | 2213 (42.2%) | 1257 (24.0%) | 5248 (100.0%) |  |
|  | yes | 21 (3.7%) | 206 (36.0%) | 214 (37.4%) | 131 (22.9%) | 572 (100.0%) |  |
| Diabetes type I |  |  |  |  |  |  | 0.425 |
|  | no | 168 (2.9%) | 1821 (31.5%) | 2413 (41.7%) | 1380 (23.9%) | 5782 (100.0%) |  |
|  | yes | 0 (0.0%) | 16 (42.1%) | 14 (36.8%) | 8 (21.1%) | 38 (100.0%) |  |
| Diabetes type II |  |  |  |  |  |  | < 0.001 |
|  | no | 152 (2.7%) | 1724 (30.7%) | 2365 (42.1%) | 1374 (24.5%) | 5615 (100.0%) |  |
|  | yes | 16 (7.8%) | 113 (55.1%) | 62 (30.2%) | 14 (6.8%) | 205 (100.0%) |  |
| Lung Disease |  |  |  |  |  |  | < 0.001 |
|  | no | 139 (2.8%) | 1544 (30.8%) | 2097 (41.8%) | 1232 (24.6%) | 5012 (100.0%) |  |
|  | yes | 29 (3.6%) | 293 (36.3%) | 330 (40.8%) | 156 (19.3%) | 808 (100.0%) |  |
| Cancer |  |  |  |  |  |  | 0.026 |
|  | no | 157 (2.9%) | 1714 (31.3%) | 2279 (41.6%) | 1329 (24.3%) | 5479 (100.0%) |  |
|  | yes | 11 (3.2%) | 123 (36.1%) | 148 (43.4%) | 59 (17.3%) | 341 (100.0%) |  |
| Stroke |  |  |  |  |  |  | < 0.001 |
|  | no | 160 (2.8%) | 1809 (31.5%) | 2397 (41.7%) | 1378 (24.0%) | 5744 (100.0%) |  |
|  | yes | 8 (10.5%) | 28 (36.8%) | 30 (39.5%) | 10 (13.2%) | 76 (100.0%) |  |
| Heart disease |  |  |  |  |  |  | < 0.001 |
|  | no | 159 (2.8%) | 1769 (31.3%) | 2353 (41.6%) | 1370 (24.2%) | 5651 (100.0%) |  |
|  | yes | 9 (5.3%) | 68 (40.2%) | 74 (43.8%) | 18 (10.7%) | 169 (100.0%) |  |
| Hypertension |  |  |  |  |  |  | < 0.001 |
|  | no | 129 (2.6%) | 1412 (28.9%) | 2085 (42.6%) | 1264 (25.8%) | 4890 (100.0%) |  |
|  | yes | 39 (4.2%) | 425 (45.7%) | 342 (36.8%) | 124 (13.3%) | 930 (100.0%) |  |
| Obesity |  |  |  |  |  |  | < 0.001 |
|  | no | 113 (2.3%) | 1381 (27.9%) | 2151 (43.5%) | 1300 (26.3%) | 4945 (100.0%) |  |
|  | yes | 55 (6.3%) | 456 (52.1%) | 276 (31.5%) | 88 (10.1%) | 875 (100.0%) |  |
| P value : Pearson's Chi Squared Test |  |  |  |  |  |  |  |

The majority of respondents (78%) were female and most (83%) were aged between 35 and 69 years. There was a relatively normal distribution of household incomes but a large proportion of the participants (62.9%, n = 3,659) were educated to degree level or higher. Participants lived across the UK including 6% in Scotland, 5% in Wales, 1% in Northern Ireland, and of those from England, 35% in London and the South-East regions. Ethnically, 95.4% of participants were white, with just 3.7% being from black and minority ethnic (BAME) backgrounds. 0.9% of respondents opted not to reveal their ethnicity. Ethnicity was not included as a covariate in statistical analyses as the numbers were too small. Similarly, non-male, non-female gendered (n = 55) participants were excluded from analysis due to limited numbers. Adults were less likely to report vigorous PA prior to the lockdown if they were female, older, had fewer educational qualifications, lower household income, lived alone, had a garden, had school-aged children and if they had obesity, hypertension, type II diabetes, lung disease, one or more ADLs and depression during the epidemic (Table 2).

**Table 2:**
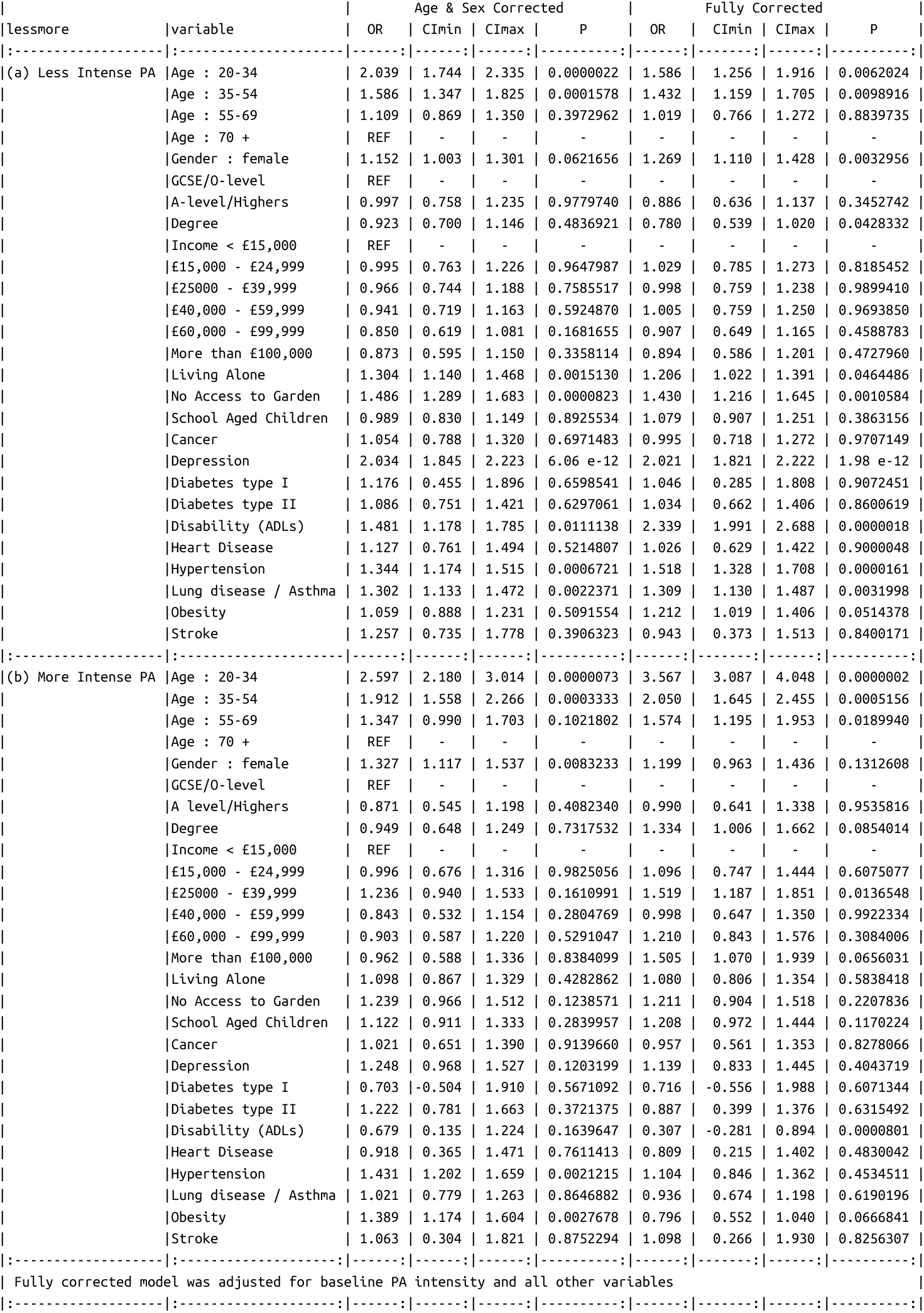
Multinomial regression: Change in PA Intensity

Approximately 37% of participants (n = 2,161) reported a change in their PA behaviours during lockdown, with 25.4% (n = 1,479) doing less and 11.7% (n = 682) doing more than before the pandemic. After correcting for baseline PA intensity, there were significantly increased odds for women (compared to men) to have started doing less intense PA under lockdown (OR 1.27, 95% CI 1.11-1.43, p = 0.003). This was also the case for people who did not have access to a garden (OR 1.43, 95% CI 1.22-1.65, p = 0.001). Older people appeared to be more likely to be doing the same intensity of PA during lockdown and compared to the group aged 70 and over, the 20-34 year olds were significantly more likely to have changed to either less (OR 1.59, 95% CI 1.26-1.92, p = 0.006) or more (OR 3.57, 95% CI 3.09-4.05, p = 0.0000002) intense PA. Decreasing age had a linear relationship to the odds of changing PA behaviours in either direction (Figure 1).

**Figure 1:**
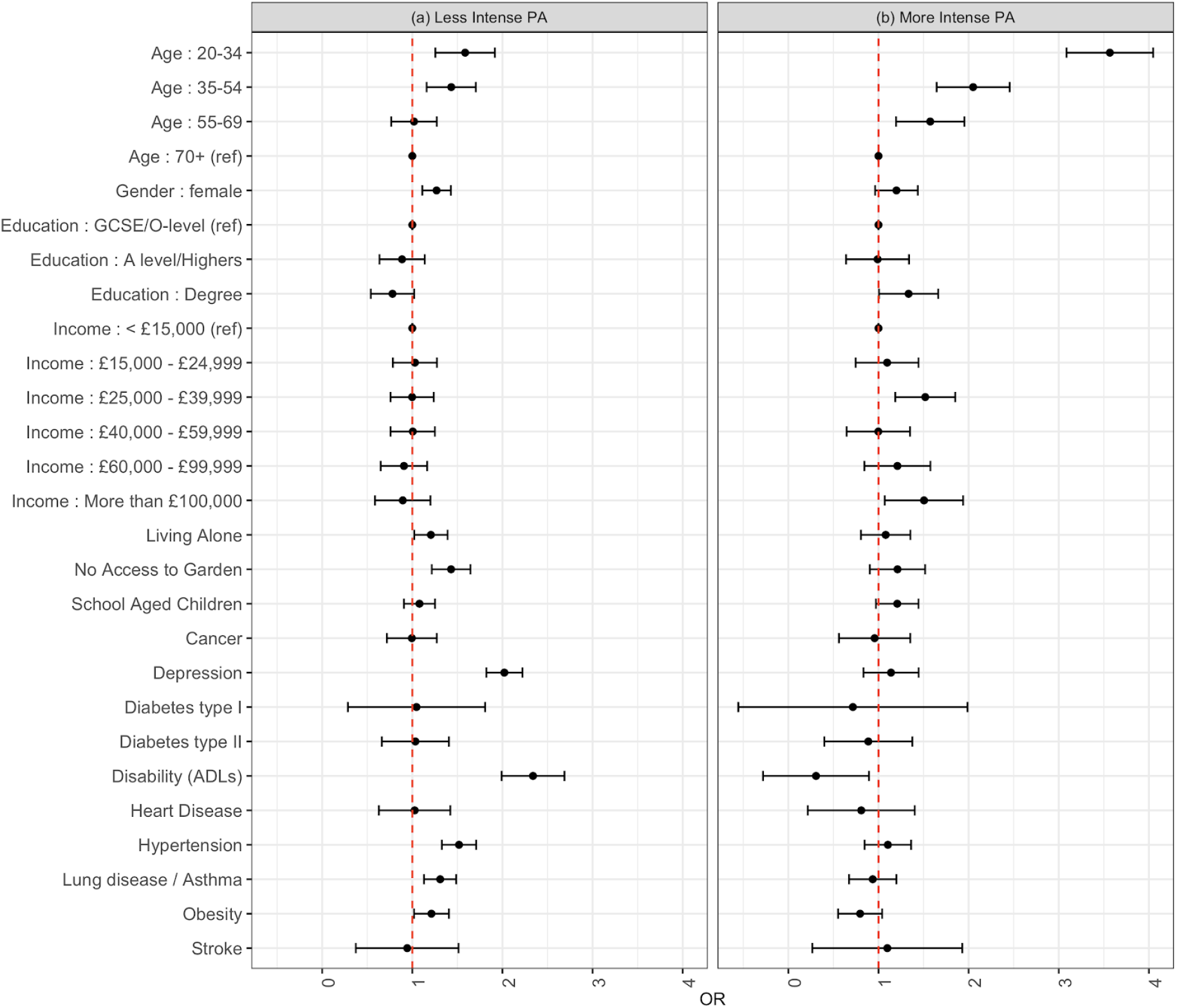
Odds ratios for having changed towards (a) less intense and (b) more intense physical activity since the UK COVID-19 lockdown began. The reference group is study participants who continued to do the same intensity of Physical Activity. All odds ratios are corrected for baseline physical activity intensity.

Lung diseases were significantly associated with increased odds of change towards doing less intense PA (OR 1.31, 95% CI 1.13-1.49, p = 0.003), which still held true in a sensitivity analysis when we did not include the additional asthma cases in the lung disease category (OR 1.306, 95% CI 1.05-1.56, p = 0.04). Hypertension (OR 1.52, 95% CI 1.33-1.71, p = 0.00002), depression (OR 2.02, 95% CI 1.82-2.22, p = 4.25 * 10^12^) and disability from one or more ADLs (OR 2.34, 95% CI 1.99-2.69, p = 1.6 * 10^−5^) were all significantly associated with change towards less intense PA behaviours (Table 2, Figure 1). All statistical testing used the group who had not changed PA intensity as the reference group.

To investigate the role of self-perceived risks on PA behaviours during lockdown, we used STM to reveal 10 topics in the 5506 survey responses which constituted the corpus of text on the coping-strategies of the study participants. The 10 key topics we identified were (T1) “Virtual Meetings/Online life”, (T2) “Buying Food, Handwashing”, (T3) “Keyworkers, NHS, essential jobs”, (T4) “Perceptions of risk to self or household members”, (T5) “Activities around the house”, (T6) “Psycho-social effects of lockdown”, (T7) “Playing games/Quality Time Together”, (T8) “Walking as part of structured routine”, (T9) “Exercise & Exercise routines” and (T10) “Children/Grandchildren, food & drink”. Representative perspectives (in the form of quotes) from the topics are provided in Supplementary Table S3. Three topics related directly to PA (T5, T8 and T9) and these PA related topics accounted for around 40% of all text content in the corpus. Topic T5 (“Activities around the house”) appeared to encompass the kind of moderate intensity PA that takes place primarily in the home and which is focussed around activities such as gardening, whilst T9 (“Exercise & Exercise routine”) appeared to refer more explicitly to exercise for fitness. T8 (“Walking as part of structured routine”) referenced the kind of mild exercise (specifically walking) that one does in the context of everyday routine such as going to the shops or work, or walking the dog. Figure 2 is a correlogram for the 10 SMT topics, which highlights how participants whose responses were classified as relating to topic T4 “Perceptions of risk to self or household members” were also likely to feature text relating to the psycho-social effects of lockdown (T9) and what we considered to be the least active of the three exercise related topics (T8). The three exercise related topics T5, T8 and T9 were largely exclusive, meaning that individual participant responses were unlikely to cover more than just one of these topics.

**Figure 2:**
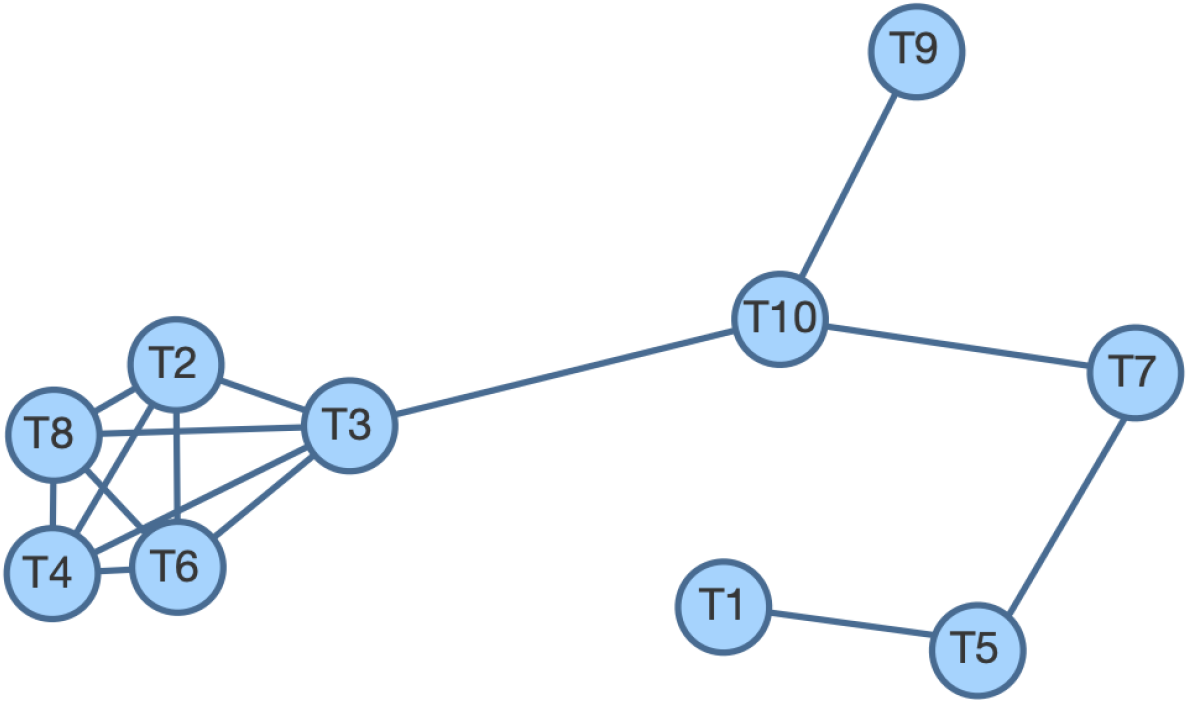
Topic Correlations in the structural topic model for the corpus of text describing coping strategies. Nodes show topics and lines show positive correlations between the topics. There was a close correlation between topic T4 “Perceptions of risk to self or household members” and topics relating to the psycho-social impacts of lockdown (topic T6), more gentle PA (topic T8) and challenges of daily life (topic T2). Those whose responses were focussed towards discussion of more intense PA (topic T9) or activities (including PA) around the house (topic T5) were less likely to also focus on T4, T6, T2 or T8. The topic correlation cutoff was 0.01.

When we performed a statistical analysis of how change in PA intensity related to coping strategies during lockdown, the STM expected text proportions revealed that perceptions of risk to self or household members (topic T4) were mentioned in 9.7% (9.0-10.5%) of responses from participants who had changed towards less intense PA during lockdown (Figure 3). This was significantly more (p = 0.00042) than the 8.2% (7.7-8.6%) of responses linked to topic T4 in participants who were doing the same intensity of PA. Topic T5 (Activities around the house) featured in a significantly lower (p = 0.00073) proportion of responses from the reduced PA group. The ETPs for topics T6 (Psycho-social effects of lockdown, P = 0.0037) and T2 (Buying food, handwashing, p = 0.019) were significantly lower in the group doing more intense PA, whilst in the same group, topic T9 (Exercise and exercise routine, p = 0.0078) had a higher ETP. The ETP for topic 10 (children, grandchildren, food & drink) were higher both for participants who changed towards more (p = 0.0268) and less (p = 0.00009) intense PA.

**Figure 3:**
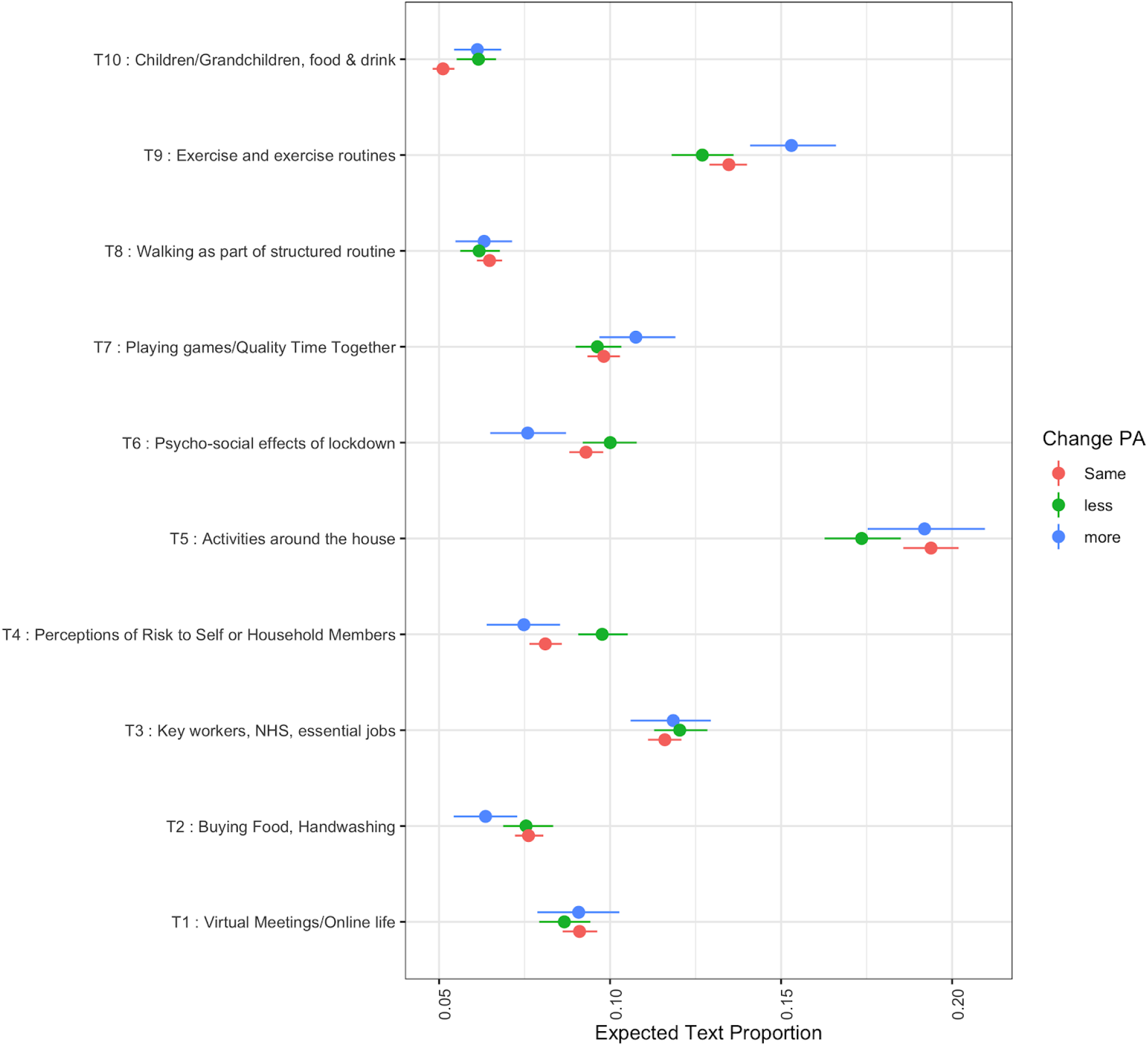
Expected text proportions in open-ended narratives on participants’ coping behaviours. Participants were asked to describe their coping behaviours during the UK COVID-19 related lockdown. Perceptions of risk to self or household members (topic T4) were mentioned in 9.7% (9.0-10.5%) of responses from participants who had changed towards less intense PA during the lockdown. This was significantly more (p = 0.00042) than the 8.2% (7.7-8.6%) of responses linked to T4 in participants who were doing the same intensity of PA. Discussion of the psychosocial effects of lockdown (T6) featured in proportionally fewer responses from people doing more intense PA (p = 0.0037).

## Discussion

In this large UK-wide survey of adults aged 20 and over we show that the majority (~60%) of the study sample succeeded in maintaining their normal PA intensity level during the study period of COVID-19 lockdown. Among those who changed their PA levels, more than twice as many people reduced their PA intensity as increased it. Adults who reported having a doctor’s diagnosis of obesity, hypertension, lung disease (including asthma), depression or at least one ADL were more likely to be doing less intensive PA compared to their activity before the epidemic. Compared to the oldest age group (70+), younger age groups were significantly more likely to have changed and to be doing either more, or indeed less intense PA since the lockdown began. Being female, living alone or being without a garden were also associated with doing less intensive PA during the study period. Importantly, we found these associations were independent from all identified confounders. We also applied ML-based text mining to open ended text data about participants’ lockdown coping strategies and found that people who expressed sentiments about personal or household risks were more likely to have exhibited a PA behaviour change towards less intense activity. This is important because individual level behaviour change is guided by both subjective and objective risks and because perceptions of risk may act as a conditioning factor in a participant’s balancing of concerns of safety, self-isolation and health during the COVID-19 lockdown.

The strengths of this study include the large population sample of adults who provided information on a wide range of demographic factors and health conditions in addition to PA behaviours before and during the COVID-19 lockdown. Our mixed methods approach allowed us to capture not only objective medical risks for COVID-19 from doctor diagnosed conditions, but also participants’ self-perceived risks which make less intensive PA more likely. We applied a recently described ML approach to the codification of topics from open-ended questions, eliminating much of the subjectivity that is usually associated with anthropological & ethnographic approaches to text-mining. Limitations of the study do exist, particularly in that this study relied on self-reported information (eg: intensity of PA, medical conditions) leaving it susceptible to response bias (e.g. imprecise recall, influence of social desirability), however we minimised this where possible, for instance by giving examples of different types of physical activity, with corresponding intensities and asking about medical conditions that were diagnosed by a physician. Whilst the ML approach we used for text mining was fully reproducible and largely autonomous, topic labels were added manually and the findings of this part of the work should be interpreted with reference to the perspectives presented in Supplementary Table S3. This study is observational and therefore causal links between the outcomes and exposures cannot be assumed. Confounders that were not included in the study or those that were misclassified may lead to residual confounding. A significant limitation is that we could not assess the role of ethnicity, which is particularly important because there is substantial evidence that there is a disproportionate effect of COVID-19 on minority ethnic groups [18,19] and because people from minority ethnic groups have worse health than the overall population, especially among those over 60 [20], The study findings are not generalisable, as with many epidemiological surveys participants were disproportionately likely to be highly educated, white and female.

The extent to which adults in the UK will revert back to their usual PA regimes once lockdown measures are relaxed is unclear, but the potential for multiple lockdowns being necessary over a protracted period could lead to prolonged periods of low PA in a substantial proportion of the population. This is concerning because it is well established that insufficient levels of PA are associated with poor mental [6] and physical [7,8] health and with premature mortality [9]. Furthermore, a reduction in PA levels for even short durations (for example a decrease in step-counts per day for two weeks) are associated with indicators of poor health including reduced insulin sensitivity, cardiorespiratory fitness, muscle mass and increased central fat [21,22]. The results of our current study suggest that the health of adults who have disabilities, depression, obesity, hypertension and lung disease may be disproportionately impacted because they are more likely to reduce the intensity of their PA. Scientists have recently published recommendations for self-isolation, including that individuals should attempt to increase their PA (even if only by a little) and to exercise every day in order to improve physical cardio-respiratory fitness in case they contract coronavirus and become severely ill [5]. This advice may be even more pertinent for those who are at higher risk of complications from underlying health conditions such as obesity and lung conditions, or for those without gardens. Our findings suggest that these sub-populations are more likely to be doing less than before the lockdown. New advice that promotes home-based exercises such as including extra daily step counts [5] and more intensive forms of PA [23] should be considered as part of any new public health guidelines for self-isolation and future lockdowns. Targeting PA health messaging to address the potential harms of subjective risks may also be key, given that those who have no known objective clinical risk in the current epidemic may change PA behaviour in light of their perception of risk, thereby driving the development of clinical risk factors and as a consequence potentially suffering more severe sequelae of SARS-CoV-2 infections during future epidemics.

To date, studies examining changes in PA before and during COVID-19 lockdown are very limited in number; this is the first study using data from the UK to examine changes in PA intensity during the COVID-19 lockdown. The results of this study are in line with recent findings from an online survey (n =1,047) of participants from across different continents, which indicate that home confinement due to COVID-19 could negatively impact participation in PA such that it was associated with a 35% reduction (equivalent to 2.45 days) in the number of days per week walking [24],

Lockdown measures due to COVID-19 are associated with a reduction in the intensity of PA in adults with obesity, hypertension, lung disease, disability and depression. Participants more frequently expressed sentiments and perspectives on risk to self or household members when they had changed towards less intense PA. Future research questions should examine how adults with and without chronic health conditions can maintain a healthy PA regime whilst adhering to lockdown restrictions.

## Data Availability

Quantitative data are available on LSHTM Data Compass https://datacompass.lshtm.ac.uk/

Data entity # 1753

Qualitative data are not publicly shared as some information could be disclosive. Access to qualitative data may be sharable and interested parties should contact the corresponding author in the first instance.

## Acknowledgements

The study team would like to thank the study participants for their contributions to the data. We are also grateful to Matthew MacGregor & Suman Golia for their contributions to the software platform and to Eleanor Martins & Esther Amon for administrative support.

## Transparency statement

This manuscript’s guarantor (ChR) declares that this manuscript is an honest, accurate, and transparent account of the study being reported; that no important aspects of the study have been omitted; and that any discrepancies from the study as originally planned (and, if relevant, registered) have been explained.

## Patient consent for publication

All participants gave consent to be involved in the study and confirmed that they understood that the data would be published in the public domain.

## Data availability statement

### Quantitative data are available (CC-BY) through LSHTM Data Compass

(https://datacompass.lshtm.ac.uk/) and have the accession number 1753. To enquire about access to the qualitative data, interested parties should contact the corresponding author in the first instance.

### Dissemination to participants and related patient and public communities

At recruitment, all participants were provided a link to our study results website. On this site we provide summary statistics and narratives on results and analysis. A link to this manuscript will be included on the project website.

### Provenance and peer review

Not commissioned. Not peer reviewed

## Supplementary Data

**Supplementary Table S1:** Correlation between previous doctor diagnosis with lung disease and mention of asthma in the chronic disease diagnosis corpus

| 576 | ----- | ----- | ----- | ----- |
| --- | --- | --- | --- | --- |
| 577 |  | No Mention Asthma | Mentioned Asthma | Total |
| 578 | No Lung Disease | 5012 (91.7%) | 453 ( 8.3%) | 5465 (100.0%) |
| 579 | Lung Disease | 130 (36.6%) | 225 (63.4%) | 355 (100.0%) |
| 580 | Total | 5142 (88.4%) | 678 (11.6%) | 5820 (100.0%) |
| 581 | ----- | ----- | ----- | ----- |
| 582 | Pearson's X-squared = 982.98, df = 4, p-value < 2.2e-16 |  |  |  |
| 583 | ----- | ----- | ----- | ----- |

**Supplementary Table S2:**
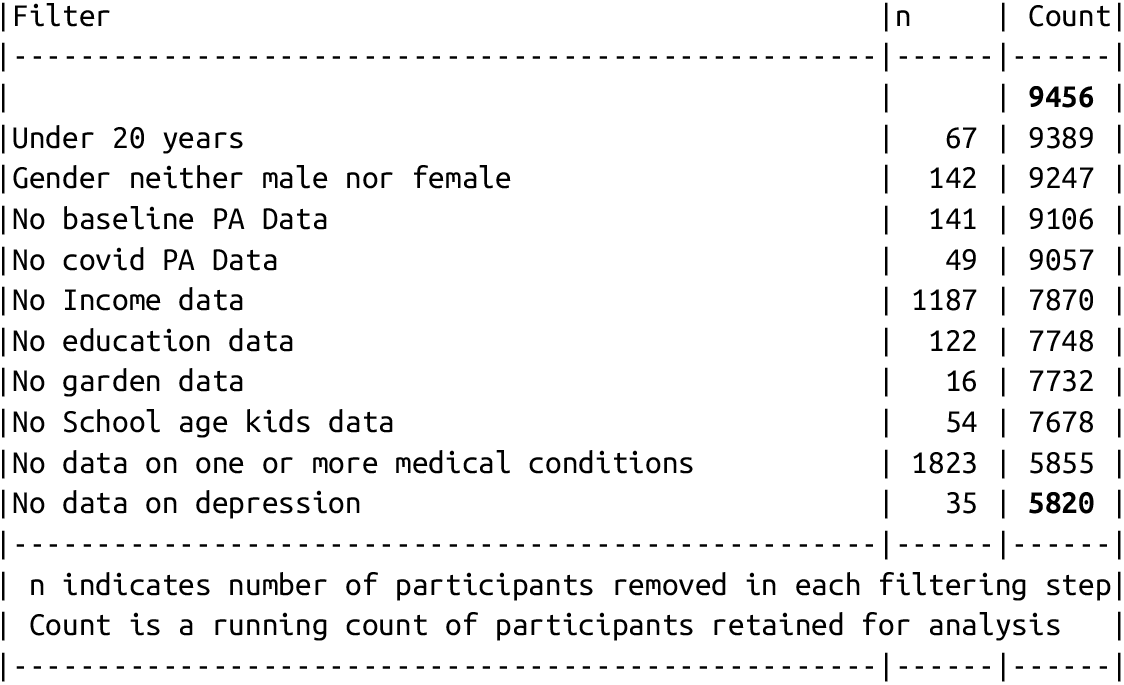
Data filtering for complete case analysis.

| 585 | Filter | n | Count |
| --- | --- | --- | --- |
| 586 | ----- | ----- | ----- |
| 587 |  |  | **9456** |
| 588 | Under 20 years | 67 | 9389 |
| 589 | Gender neither male nor female | 142 | 9247 |
| 590 | No baseline PA Data | 141 | 9106 |
| 591 | No covid PA Data | 49 | 9057 |
| 592 | No Income data | 1187 | 7870 |
| 593 | No education data | 122 | 7748 |
| 594 | No garden data | 16 | 7732 |
| 595 | No School age kids data | 54 | 7678 |
| 596 | No data on one or more medical conditions | 1823 | 5855 |
| 597 | No data on depression | 35 | **5820** |
| 598 | ----- | ----- | ----- |
| 599 | n indicates number of participants removed in each filtering step |  |  |
| 600 | Count is a running count of participants retained for analysis |  |  |
| 601 | ----- | ----- | ----- |

**Supplementary Table S3:** Topic Perspectives (Exemplar Quotes) from text corpus on coping behaviour during lockdown.

| **T1 : Virtual Meetings/Online life / Social and work life online** |
| --- |
| Set up a church community group with virtual services. Set up several Wats app groups and exchanged funny jokes and videos |
| Daily walks. Online painting tuition. Zoom meetings with my choir. Zoom meetings with my sisters. Zoom meetings with my Slimming group. Messenger video chats<br>Texting, watts app, and Facebook |
| Making funny videos sharing them. Family quizzes and singing on social media. WhatsApp groups of friends sharing memories and photos. Video calls to and from my family and friends daily. |
| Joining community and street Corvid-19 support group. Organising and helping vulnerable groups. Including various whatsapp groups. Sharing information on help and support for the community. Researching and reading medical/Science journals<br>But now ill with Corvid-19. |
| Making more calls by phone. Making/receiving emails. Playing Bridge on line. Viewing seminars on line. Sharing quizzes on line. |
| Walking/birdwatching. Video chats. Video meetings. Tv/films. Sharing memes on social media |
| Gardening, online quizzes and social meetings via zoom, art work, film and photography, telephone counselling, support from a parenting support group. |
| Set up community FaceBook page for surrounding streets. Participated in local "Quiz Nights" online using FB and Zoom. Avoiding TV news. |
| I have continued with my Zumba and Pilates via Zoom. I have had virtual meetings with friends. I'm have emailed. have talked to my family on the phone and used FaceTime to speak to my children and grandchildren. I have painted rooms and gardened plus reading doing crosswords and puzzles. |
| Email, Skype, Messenger, landline phone |
| **T2 : Buying Food, Handwashing / Shopping whilst adhering to guidance** |
| I normally visit people who are not able to get out. I have been calling them instead.<br>I have been making laundry bags for frontline staff, so that they can put their scrubs in the bags and the bags into the washing machine without touching the scrubs again.<br>Joined z local volunteer group to collect prescriptions/do shopping for those being shielded.<br>More cleaning than usual.<br>Shopping |
| Prepared grocery shopping and buying more items at supermarket. Other shopping i order online. Use alcohol bssed handwash, wipes and gel. Go out much less. Take my own shopping bags and wash them at home. Antibac handles, steering wheel and hands regularly. I get all of my sisters shopping as she has a compromised immune system. |
| Tried to ensure weeks worth of food in house when I ship to minimise going out. Try to go out at quieter times. Avoid narrow alleys etc where social distance impossible. Only go to shops with good social distance measures. |
| Asking people to help with shopping. Changing our birth plan because parents can't visit and help |
| Keep in contact with people virtually. Social distance. Wash hands |
| Avoid contact with people.<br>Not taking any public transport.<br>Using face masks when I go to buy food |
| I am a key worker in that I deliver eggs to local shops. Customers have been very good and helped with keeping a distance from me.<br>I try only to leave the farm either to deliver eggs or to deliver food to my elderly mother. |
| Wearing mask & gloves while shopping. Keeping my distance from others (even in the street), washing hands regularly & thoroughly. |
| Not leaving the house except a short walk, for a weekly food shop and to deliver food to elderly parents. |
| Shopping for elderly parents and neighbours. Volunteer for local Age UK. Working as normal. Generally still very busy |
| **T3 : Key workers, NHS, essential jobs** |
| I am classed as a key worker so have still been going to work .I have only left my home for essential reasons. |
| Key worker so still working |
| I work in a Hall of residence so have been going in to work. During weekends I've been having a clear out at home. |
| Staying at home<br>Going to work with incorrect PPE, leaving patients at home where available |
| Still going to work as an infection control nurse and my daughter has been attending school for childcare reasons as I am single parent |
| Keeping busy working from home and doing jobs around the house at weekends. |
| Going to work I'm a key worker. Nurse in a prison. |
| Staying at home, working online, in garden, jobs around house, starting my MSc dissertation, seeing my horse. |
| As a key worker I have been attending work therefore the predominant impact has been in evenings and weekends. Gardening and diy have therefore become the norm. |
| Staying at home, working from home, doing the garden and odd jobs. |
| **T4 : Perceptions of Risk to Self or Household Members** |
| I'd lockdown before then as my husband suffers from Parkinson's Disease & I wanted to protect him. I organised milk delivery & a fortnightly supermarket delivery. I made sure I had frozen fruit & veg, pulses & nuts stocked ( we are vegetarian). I paid a newspaper bill in advance. |
| Working, a lot, including caring for Covid+ patients. Then I was ordered into lockdown due to inability to care for me if I get sick (medical conditions) which I've been struggling with as I feel I've abandoned my colleagues |
| I have been semi-retired for the past 2 years so already had established routines prior to lockdown. Lockdown has not had a significant impact as I am fortunate enough to have a garden and I keep poultry |
| I live on a boat. I have not been out other than hospital for cancer treatment. Husband done everything. Not feeling too isolated. Now have access to food delivery. |
| I am a teacher who is 9 months pregnant and I currently have an 19 month old son. I started isolating when they classed pregnant women as vulnerable. My partner continued working as he is in the energy industry but has begun self isolation last week as the baby is due in 1 weeks time. We have been very lucky to have such nice weather recently as we have spent most days in our small garden. When I first started isolation we had no garden toys, but luckily our family and friends donated some to keep our son busy. The hardest part has been keeping our toddler occupied. I have done lots of cleaning!! |
| I've got a huge garden that needs loads of restoration. I put the infrastructure in last year including a kitchen garden. I've loads of seeds, two polytunnels plus a small flock of ex battery hens. Plenty to keep me busy. |
| I've been in the house since 24th Feb, cos I had Covid 19 then, and was ill for 5 weeks. I spend time with my husband; I talk to friends and family on line; I do 'colouring in, and I watch a lot of TV.<br>I also spend a lot of time trying to get supermarket deliveries for us and for my 87 year old mum, who is 350 miles away! |
| I don't feel I've had to cope, I've found it easy. |
| We have a garden and surrounded by farmland so naturally isolated. We spend time in the garden, care for our pets (on property), try and homeschool my children and maintain housekeeping routines. We have also watched a lot of children's films which are up lifting |
| We had been isolating for two weeks prior to this with a vulnerable disabled daughter. Working from home and access to private outside space with delivery of food has made it easy. |
| **T5 : Activities around the house** |
| Meditating, reading, jigsaw puzzles, sewing, knitting, tv films, gardening, walking the dog, family tree |
| Sewing, knitting, baking, gardening, reading, watching tv, listening to the radio. |
| Online yoga. Reading Gardening Cleaning house. Walking dog Streaming theatre shows Watching films Craft activities |
| Walking, gardening, reading, texting, phoning, sewing, knitting, watching television, cooking. |
| Gardening, listening to music, watching films, doing puzzles, reading |
| Reading, watching TV, watching films, cooking, tidying cupboards, gardening, beauty treatments, music |
| Decorating, gardening, cleaning, sorting garage, wardrobes, cupboards, exercising, watching tv and reading, cooking |
| Bible study gardening journaling reading drawing exercising cooking box sets cleaning |
| Housework, gardening, walking the dog, reading, knitting, social media, Netflix, identifying birdsong, citizen science entries on ispot. |
| Puzzles, gardening, internet, listening to music, watching films. |
| **T6 : Psycho-social effects of lockdown** |
| I like this way of life. I miss certain things but love the way nature is getting a break.<br>I wish we would stay like this forever with a limit on our carbon consumption and a carbon currency used. I guess thinking about these things is helping me cope, but it's not lockdown that's the problem, it's the thought of people 'returning to normal'. |
| I am not coping well. I was just emerging from a depressive episode that had lasted six months. Now I'm feeling much worse and would rather die than live like this. So many of the events that I had booked, which were what was giving me a reason to look forward to and keep going, have now been cancelled. There's nothing to look forward to and being stuck in doors with no gym or friends to see is miserable. This life is not worth living. |
| I am not coping since I have Asperger's syndrome which makes me terribly anxious. I talk to my wife who is coping better. I feel tremendous resentment at the strategy some want to pursue. Repeated lockdowns until end 2021!?!?! I repeat REPEATED, PROLONGED LOCKDOWNS ARE BARBARIC AND UNACCEPTABLE. |
| I have 6 yr old grandson living with me, he is a difficult child, so this makes coping more difficult. I would do better coping on my own. |
| Nothing different, its just more peaceful while I read, meditate and learn |
| I live by myself, in a City 30 miles from my nearest family (sister). I don't see friends very often. Apart from not being able to go out in my car, nothing has changed that much. |
| I don't find it particularly difficult; nothing has happened to me/my family yet, and I do not tend to worry ahead. I think that it is a fascinating time, in a way, and observing the social/sociological/psychological side of it is rather exciting. I would probably miss going out, but I currently have leg injury and cannot walk anyway; that is quite annoying. What I worry about is when will I be able to visit/see my family (parents/brother) - they are overseas (I am Polish). But, as I have no impact on it, I do not think about this too much. (This is, actually, quite a prompting question - why assume it is automatically 'difficult'? There may be a variety of reactions) |
| Doing exactly the same things as I did before. Hasn't changed my life much. |
| no change to my life, just can't go to the pub |
| I am alone anyway. So in a perverse way, the fact that others now have to live simply and quietly by themselves is a strange comfort.<br>Been involved in NHS all my adult life (ex husband a Doctor) In the past, was sad that I had wasted my life caring so much. Now feel proud to have carried that torch of humanity, and see it blaze in this time of uncertainty. It is all that really counts in a life lived. |
| **T7 : Playing games/Quality Time Together** |
| Playing guitar<br>Enjoying family movies together<br>riding bike / running |
| Having projects to do in the house & garden (re-upholstered an ottoman, lifted paving slabs, painted fences) catching up on incomplete tasks or tasks not yet got round to (tidying the cupboards etc).<br>Playing board games and video games with my daughter. Baking and cooking with my daughter. |
| Taking time to relax, watching films to distract me.<br>Spending time cuddling my cats. |
| Spending time with my husband. Learning to cut his hair. started exercising, yoga, spending time in my garden. Speaking and video calling friends and family. Reading. Watching tv. Learning to cook new things. Clearing up in the house. Playing bridge online. Doing brain activities. Playing games. |
| Playing computer games a lot. Worrying about long term finances. Making lists of craft projects. Playing musical instruments |
| Having a list of tasks that I have wanted to do for a long time but never had the unlimited time to complete. Like decluttering the house garden and spending time spoiling my family. |
| Decorating. Spending time playing board games. Camping in the garden. Doing the jobs I've been meaning to. Writing letters. Walking my dogs. Riding my bike |
| I have written a list of tasks in the house and garden which we are steadily completing. I'm also making a quilt, a crochet blanket for my granddaughter. I play the piano, read and take the dog out for long walks with my husband. |
| Spending quality time with my partner. Chatting (at distance) with a neighbor.<br>Calling and texting friends.<br>Watching movies, series, studying playing pc games |
| Communicating with friends and family through phone calls and messages. Doing tasks with my children. Spending time in our private garden. Family time playing games, making fun videos etc. |
| Spending time with the children playing games and helping with homework. Going on bike rides and exercising at home. Making the most of our beautiful local area |
| **T8 : Walking as part of structured routine** |
| following the rules, and knowing that each day that passes is one day nearer it all being over |
| Started following Joe Wicks' exercises every morning. Shopping twice a week (instead of 6 times) and getting bits for neighbours. |
| Having a structure to the day. A one hour walk to exercise our dog. |
| I walk two dogs 5 days a week for one hour each walk. Read books more. Gardening |
| I am an NHS key worker. I treat cancer patients. I have 3 days off per week. I cycle for one hour a day avoiding people, on days off. I try and walk around the hospital every day when at work. I shop once a week. I stay at home the rest of the time. I study, I read, I watch the news bulletins. |
| going for a walk every day |
| Luckily we have a dog and a garden. We walk the dog morning and evening for about half an hour. We do a thirty minute exercise class every day. I cook fresh food every day and we are in contact with friends who we miss seeing. We Zoom with family and friends every week and keep up to date on both radio and tv with a lighthearted movie every so often to lift the mood. |
| Structuring each day, so that it is at least reasonably productive and always includes an interaction with someone. Making sure that every day I eat healthily and go for a long walk. Keeping up to date with virus news, but not looking more than twice a day, as it could lead to excessive concern. |
| I walk my dog once a day. |
| Exercise everyday. Preferably early morning as I do walk long distances and later in the day as my wife doesn't like to go out on her own.<br>Im a musician so I practice 2-3 hours daily.<br>Clean house and social media to keep in touch and check the news. |
| **T9 : Exercise and exercise routines** |
| Keeping a fixed regular routine, exercise, meditation, keeping in touch with friends and family |
| Keeping in touch with friends and family, exercising, trying to keep routine. |
| Keeping in touch with friends and family, exercising, chores, trying to keep myself busy |
| Keeping in touch with friends and family, keeping a routine and doing creative things to keep entertained. |
| Routine, exercise, healthy diet, meditation, talking to friends, getting informed, |
| Exercise<br>Keeping in touch with family and friends<br>Working<br>Trying to eat more healthily |
| Volunteering. Setting daily targets. Keeping in touch with friends and family. Counting my blessings. Not obsessing about news reports. |
| Keeping informed, keeping busy, keeping active |
| Contact with colleagues<br>Exercise online class<br>Keeping some routine |
| Exercising, getting fresh air, mindfulness, trying to eat & sleep properly, keeping in touch with friends & colleagues. |
| **T10 : Children/Grandchildren, food & drink / Daily routines** |
| Talking about it, not putting any pressure on myself or my family (recognising this is abnormal), sleeping more, drinking more alcohol, letting the kids camp together each night, staying up late, eating better - cooking from scratch every night with what we have, being grateful for the local community |
| Focusing on giving my children a stress free time. |
| Trying not to alter my routine - going to bed and getting up at the same time<br>Trying to keep busy with studying<br>Trying not to overburden myself with the pressure to 'use' this time well<br>Making more time to talk to friends<br>Keeping in touch with my partner and sending gifts<br>Playing games and watching TV with family<br>Exercising, probably more than usual<br>Eating well, probably better than usual |
| Home deliveries keeping contact with family watching TV with my partner, cleaning everything I can planning meals doing art painting trying to relax helping others relax. Making sure children and grandchildren are OK by messaging. Trying to watch comedy as much as possible drinking tea keeping up to date trying not to panic or cause panic to others. |
| Learning new hobbies for achievement of goals.<br>Communicating with others virtually.<br>Keeping routines including sleep and mealtimes.<br>Research.<br>Making time for R&R.<br>Planning for the future.<br>Checking in on others in challenging situations.<br>Eating well with a varied, nutritious diet.<br>Clearly following government and infection control guidance. |
| Exercising at home, creating a better wfh environment, learning how to talk to friends/family via video, appreciating free time, trying not to feel overwhelmed but learning how to combat those feelings |
| Eating comfort food and drinking alcohol more than usual. |
| Drinking... lots of drinking |
| Consuming more alcohol than normal, stress eating |
| Planning days for my young children, trying to look after my patients remotely, talking to friends, running online yoga classes, drinking gin, eating chocolate, helping neighbours and gardening |
| **Note : These texts and quotes are presented in the original form, which may include spelling and grammatical errors, as well as the use of language that some may find offensive. These quotes are perspectives of the study participants and do not reflect the opinions of the study authors.** |

**Supplementary Figure S1:**
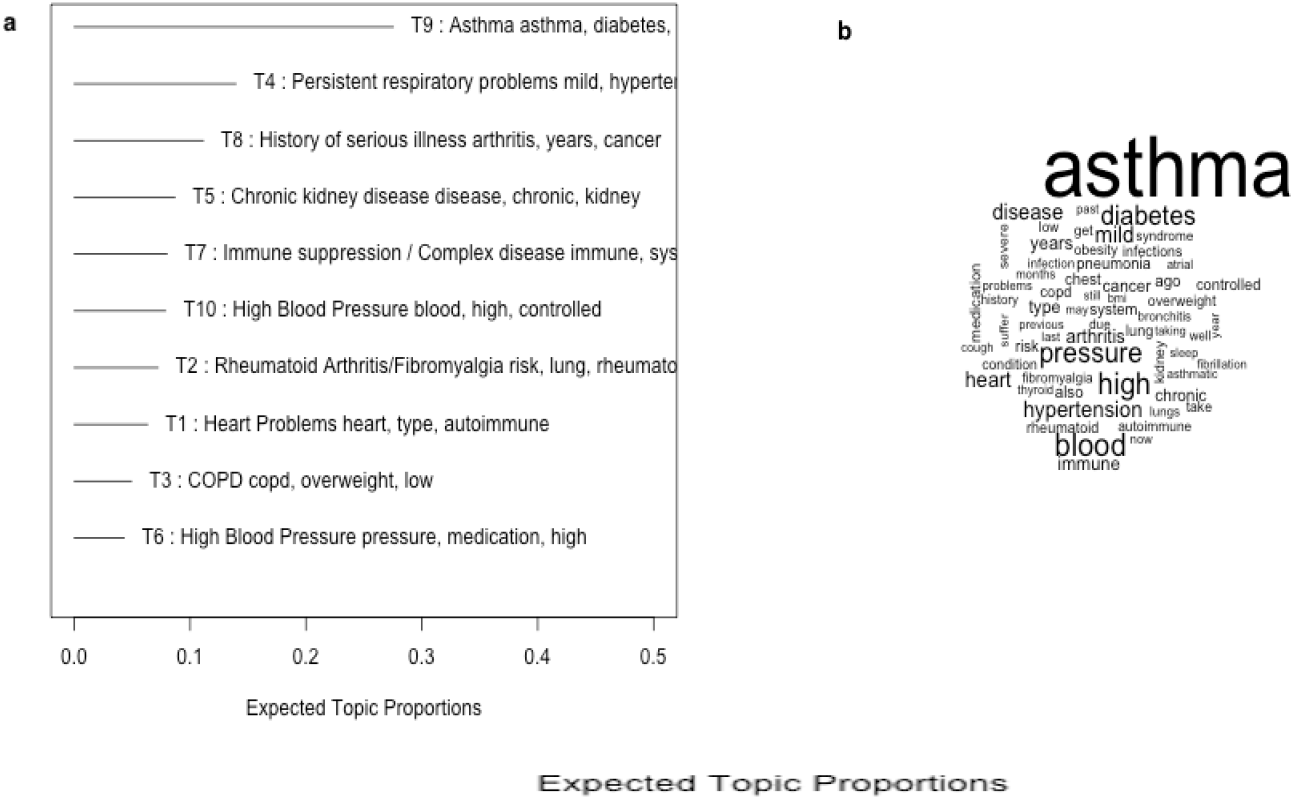
Topic Proportions (a) and word frequency cloud (b) for STM of chronic disease diagnosis free-text data. Asthma was the most frequently mentioned word and an asthma related topic was the most prevalent among the corpus of text.

